# Long-term Metabolic Side Effects of Second-Generation Antipsychotics in Chinese Patients with Schizophrenia: A Within-Subject Approach with modelling of dosage effects

**DOI:** 10.1101/2024.03.04.24303695

**Authors:** Kenneth C.Y. Wong, Perry B.M. Leung, Benedict K.W. Lee, Pak C. Sham, Simon S.Y. Lui, Hon-Cheong So

## Abstract

**Background:** Second-generation antipsychotics (SGAs) are commonly used to treat schizophrenia (SCZ), but SGAs may differ in the severity of side effects. Previous observational studies had limitations like failing to account for confounding factors and short follow-up periods. This study compared the long-term metabolic and anthropometric side effects of seven second-generation antipsychotics (SGAs) in a Chinese schizophrenia population, using a within-subject approach to reduce risk of confounding.

**Methods:** Longitudinal data on SGA prescriptions, concomitant medications, fasting blood glucose, lipid profiles, and BMI were collected from 767 schizophrenia patients, with follow-up up to 18.7 years (median ∼6.2 years). Linear mixed models estimated the effects of SGAs, as binary predictors and by dosage, on metabolic profiles.

**Results:** When considering SGAs as binary predictors, clozapine and olanzapine were associated with the most substantial worsening of lipid profiles and BMI, while clozapine alone showed a significant increase in blood glucose. Amisulpride, paliperidone, and quetiapine worsened lipid profiles and increased BMI. Conversely, aripiprazole improved lipid profiles but slightly increased BMI. Examining dosage effects showed consistent results overall. At minimum effective doses, clozapine had the most severe metabolic side effects, followed by olanzapine. Risperidone and aripiprazole had the least metabolic impact, with aripiprazole significantly lowering lipids.

**Conclusions:** This study clarified the long-term, dose-dependent metabolic and anthropometric effects of different SGAs in Chinese schizophrenia patients. Our findings may inform clinicians and SCZ patients of SGA choices.

## 1. Introduction

Antipsychotic medications are mainstream treatments for schizophrenia (SCZ)[1]. Second-generation antipsychotic medications (SGAs) comprise pharmacological agents with different chemical structures and receptor affinities and have been widely used in treating psychosis, including first-episode SCZ patients[2-4]. Compared to first-generation antipsychotic medications (FGAs)[5], SGAs are associated with fewer extrapyramidal side effects[6]. However, SGAs can accompany anthropometric and metabolic side effects, including weight gain, diabetes mellitus and dyslipidemia[7-10], resulting in metabolic syndrome and cardiovascular diseases[11-13]. SCZ is associated with an average of around 15 years of potential life lost, mainly due to increased mortality caused by physical comorbidities, in particular cardiovascular disorders [14, 15].

Recent network meta-analysis of RCTs compared the efficacy and tolerability of different antipsychotics, and concluded that SGAs differed markedly in metabolic side effects[7, 16]. Despite these empirical evidence[7, 8, 17, 18], previous research on side effects of SGAs has several limitations, including modest sample size[19, 20], lacking baseline (pre-SGA prescription) metabolic data[6], failing to discern the causal relationship between SGAs and metabolic side effects (owing to confounding)[19-22], short duration of longitudinal observations (ranged from weeks to around one year)[7, 9, 10], and failing to account for concomitant prescriptions[7, 10, 21].

We note that a number of RCTs (and meta-analyses of RCTs) have reported metabolic side effects of SGAs, and RCTs are generally regarded as free of confounding. However, RCTs may have shortcomings. One important limitation is that RCTs are usually of short duration. For example, a recent meta-analysis on “long-term” metabolic side effects of antipsychotics had a median follow-up of only less than a year (45 weeks), and the dropout rate was high (up to 42%); many RCTs had even shorter follow-ups[16]. Besides, RCTs are primarily intended to study the efficacy of SGAs and are often conducted in selected subjects without physical comorbidities and concomitant medications. As such, the generalizability of RCT findings to the real-world setting may be a concern. In recent years, there has been growing recognition of the need to complement RCTs with real-world observational studies[23]. Moreover, heterogeneity between RCTs may further limit the generalizability of findings from RCT meta-analyses. Notably, many previous studies in SGAs’ effects were conducted in European populations, and rarely on Asian/Chinese settings. Given the difference in baseline BMI and metabolic profiles between Asian and European populations[24-27], it is valuable to study SGAs’ effects using a Chinese sample.

Careful design and novel analytic approaches can mitigate confounding, including unmeasured confounding, in observational studies. Here we disentangle the between-subject estimate from the within-subject estimate to reduce confounding bias when estimating associations between SGAs and metabolic measures[28]. In a hypothetical illustration that we set up in Figure 1, when looking at the between-subject estimate, a drug for treating diabetes may seemingly be associated with higher glucose levels. Yet if we look at the within-subject estimate, the same drug actually reduces an individual’s glucose levels. This issue of “confounding by indication/contra-indication”[29] can be addressed using advanced statistical modelling, such as the hybrid linear mixed model (LMM) (employed in this study) [30], which can be applied to longitudinal data for disentangling the within-subject estimate from the between-subject estimate of the effects of drugs[31-33]. By estimating the “within-subject changes”, we could account for unknown or unmeasured time-invariant confounders.

**Fig. 1.**
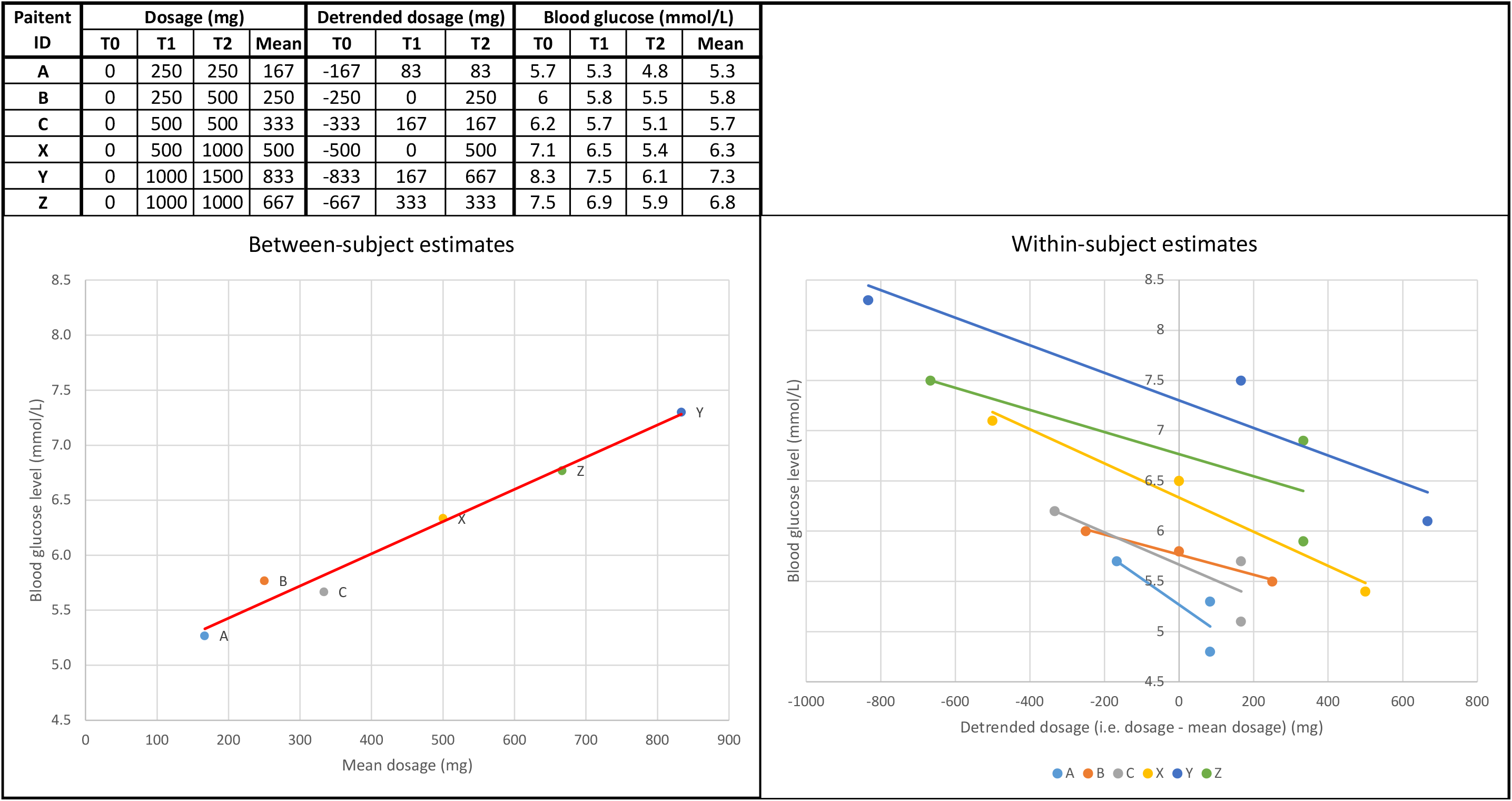
A hypothetical example to illustrate the concept of the between-subject and the within-subject estimates. Blood glucose levels were measured at times T0, T1, and T2. Patients A, B and C with prediabetes were prescribed lower dosages of diabetes drugs at time T1 and T2, respectively; while patients X, Y and Z with diabetes were prescribed higher dosages of diabetes drugs at time T1 and T2, respectively. The between-subject estimates (left) misinterpret that higher dosages of diabetes drugs are associated with higher blood glucose levels (i.e., confounding by indication). This confounding is minimized in the within-subject estimates (right) which correctly indicate a higher dosage of diabetes drugs is associated with lower blood glucose levels.

Previous research mainly studied the differences in the severity of metabolic side effects between SCZ patients exposed to SGAs and controls, and seldom investigated the dose-dependent side effects[7, 8, 10, 34]. Clarifying dose-dependent side effects of SGAs on SCZ patients is important for better prescribing practice.

To the best of our knowledge, this is the first long-term (median follow-up > 5 years) longitudinal study on the (quantitative) anthropometric and metabolic side effects of SGAs, and the first study of such in the Chinese population setting. This study attempted to apply the hybrid LMM to a longitudinal cohort of SCZ patients who received SGAs for a period of 0.4 to 18.7 years (median ∼6.2 years). We aimed to compare long-term side effects on anthropometric and metabolic parameters between different SGAs in Chinese SCZ patients.

This study has several unique strengths. Firstly, the use of within-subject estimates could substantially reduce the risk of confounding by indication/contra-indication. Second, we investigated the effects of SGA dosage on metabolic profiles, which have been seldom studied previously. Thirdly, the median follow-up duration for our sample was 6.2 years, with a maximum duration of 18.7 years, which was longer than most RCTs or observational studies in this area. Fourth, we employed linear mixed models to account for longitudinal changes in metabolic profiles, instead of using cross-sectional models. Fifth, the study controlled for the use of other drugs, ensuring that the observed effects are due to the SGAs instead of being confounded by other medications. Sixth, this study involved data of up to 27,723 metabolic measures and 192,152 prescription records, covering 767 SCZ patients. To our knowledge, this study was the largest longitudinal observational studies to date for examining a wide range of metabolic abnormalities in Chinese SCZ patients.

## 2. Method

### 2.1 Longitudinal dataset

SCZ participants were recruited from a research-oriented clinical programme at Castle Peak Hospital (CPH) Hong Kong in 2009-2021[35]. The inclusion criteria were (1) aged >18, (2) ethnic Chinese, (3) ICD-10 diagnosis of SCZ or schizoaffective disorders, (4) received SGAs, (5) having pre-SGA baseline measures of BMI, BG and lipid profiles, and (6) having >1 post-SGA measures of BMI, BG and lipid profiles. Exclusion criteria included (1) a known history of metabolic disorders (diabetes mellitus or dyslipidemia) before SGA prescriptions, and (2) did not have psychiatric follow-up as of March 2021. Written informed consent was obtained from all participants.

Based on the existing cohort of 773 SCZ participants, we reviewed the computerized clinical records to retrieve all the available measures of BMI, BG and lipid profiles for each participant, from service entry to assessment endpoint. According to the territory-based guidelines, all participants who received SGA are recommended to have baseline (pre-SGA) measures for the following items, namely BMI, BG, fasting high- and low-density lipoproteins (LDL and HDL), triglycerides (TG), and total cholesterol (TC). These measurements were repeated annually until the cessation of SGA medications.

Moreover, we retrieved each participant’s full prescription records during the entire observation period. The dosage of SGA (mg), and use of antidepressants, antidiabetic drugs and lipid-lowering drugs were all recorded.

### 2.2 Data cleaning and pre-processing

We excluded six subjects with duplications and inconsistent clinical parameters, resulting in the final dataset of 767 participants with SCZ. A total of 192,152 prescription records were retrieved, among which 19,316 records were included in the final analysis as they were dated close to the metabolic measures (see Supplementary Table S2). The measures for BMI, as well as the fasting blood collection (i.e., BG, LDL, HDL, TG, TC), could be conducted at different time points. Therefore, we compiled three datasets, i.e., lipid profile (TC, HDL, LDL, TG), BG and BMI, which contained 4,051, 4,076, and 7,598 metabolic measures respectively. After removing those with missing values, the three datasets contained 4,050 measures for TC, 3,917 measures for HDL, 4,037 measures for LDL, 4,045 measures for TG, 4,076 measures for BG, and 7,598 measures for BMI.

Before entering the data into the hybrid LMM models, we removed those measures having >6 SD beyond the group mean as outliers. Specifically, we removed 2 measures of TC, HDL, BMI, and 11 measures for TG, and 27 measures for BG as outliers. No outlier was observed for LDL. Consequently, 4048 measures for TC, 3917 measures for HDL, 4035 measures for LDL, 4034 measures for TG, 4049 measures for BG, and 7596 measures for BMI were retained for analysis. We transformed these variables of interest using natural log transformation and conducted a QQ plot (see Supplementary Figure S1).

### 2.3 Variable selection for modelling

Regarding the data of prescription records, a few types of SGAs had only been prescribed to a small number of SCZ participants throughout the observation period. To build reliable models, we examined the effects of seven SGAs, i.e., clozapine, olanzapine, aripiprazole, amisulpride, paliperidone, risperidone, and quetiapine, which were prescribed in a sufficient number of patients (>30). Long-acting injectable and oral preparations were analysed in the same way, after dose conversion[36]. Given other psychotropic drugs can affect BMI, BG and lipid profiles, we accounted for the effects of concomitant prescriptions, including lithium, valproate, sertraline, and citalopram on anthropometric and metabolic measures. Besides, we accounted for the effects of metformin, simvastatin, and atorvastatin which were commenced during the observation period, similar to prior studies[37-40]. These concomitant prescriptions were used in >30 participants in our cohort.

Age, gender, years of education and treatment duration were included as covariates. To address multicollinearity, we estimated the correlations between the variables of interest and covariates. As shown in Supplementary Figure S2, no significant correlation had been found, except for simvastatin and metformin. Given that these two drugs were included as covariates, and our study did not estimate the effect size of these two drugs, the multicollinearity as such would unlikely introduce bias to our findings.

The authors assert that all procedures contributing to this work comply with the ethical standards of the relevant national and institutional committees on human experimentation and with the Helsinki Declaration of 1975, as revised in 2013. All procedures involving human subjects/patients were approved by the ethics committees of the New Territories West Cluster (reference number: NTWC/CREC/823/10; NTWC/CREC/1293/14) and the Chinese University of Hong Kong and the New Territories East Cluster (reference number: 2016.559).

### 2.4 Considering the “side effect latency period”

We postulated that SGA-induced metabolic side effects would take time to emerge. In this study, the “side effect latency period” was defined as the time lapse between the initiation of SGA medication and observed changes in our variables of interest. We then tested the linear mixed models with different latency periods (2, 4, 7, 14, 21, 28, and 35 days) to determine the optimal model with the minimum Akaike Information Criterion (AIC). The AIC values were plotted against the latency period in Supplementary Table S1. The optimal latency period ranged from 7 to 35 days for TC and HDL measures. The AIC values for models with a 21-day latency period showed consistently close-to-minimal values and were chosen for all subsequent models in this study.

### 2.5 The between-subject and within-subject estimate of SGAs on variables of interest

We employed the hybrid LMM to investigate the association of “time-varying” variables with outcome variables, and to disentangle the within-subject estimate from the between-subject estimate[22]. In hybrid LMM, the between-subject component used the expected value of the risk factor (i.e., mean) as the independent variable, while the within-subject (time-varying) component reflected the differences between the subject’s risk factor levels (here the risk factor is SGA prescription [coded as 0/1] or the dosage of SGA) and their expected levels at different time points[30] (i.e., detrending of subject’s values). This approach addressed the issue of “confounding by indication/contra-indication”, and disentangled the between-subject estimate from the within-subject estimate (see Figure 1). It is noteworthy that LMM can accommodate imbalanced longitudinal data (i.e., irregular measurements and different follow-up durations for each person) and missing values.

When disentangling the relationship between a time-varying variable (such as clozapine doses taken by a subject *i* – *dose. CLOZAPINE*_(*t-*21),*i*_ ) and the corresponding metabolic level measured at time *t* – *BMI*_*ti*_, the hybrid LMM took two components into consideration, i.e., the between-subject variable (*bz_dose. CLOZAPINE*_*i*_) and the within-subject variable (*wz_dose. CLOZAPINE*_(*t-*21),*i*_). We incorporated the optimal latency period of 21 days into these variables, and used the dose taken 21 days before time *t* (i.e., *dose. CLOZAPINE*_(*t-*21),*i*_ ), rather than dose taken at time *t* (i.e., *dose. CLOZAPINE*_*ti*_ ), as the time-varying variable. In our study, the between-subject *bz_dose. CLOZAPINE*_*i*_ and within-subject *wz_dose. CLOZAPINE*_(*t-*21),*i*_ variables were specified in the hybrid LMM as described below, following the methodology proposed by Curran and Bauer [22]:

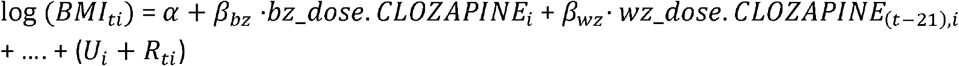

{*U*_*i*_ is a random effect and *R*_*ti*_ is the time- and subject-specific residual. *dose. CLOZAPINE*_(*t-*21),*i*_ was parsed into *bz_dose. CLOZAPINE*_*i*_ and *wz_dose. CLOZAPINE*_(*t-*21),*i*_, using the following linear regression model:

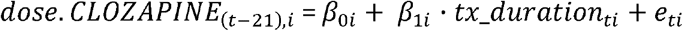

In this formula, the covariate *tx_duration*_*ti*_ represents the treatment duration at time *t* for the subject *i*, where the corresponding metabolic levels were measured at time *t*. If there is no medication before the first metabolic measurement at *t*=0 (i.e., *tx_duration*_*ti*_= 0), *dose. CLOZAPINE*_(*t-*21),*i*_ will be zero. Intercept *β*_0*i*_ reflects the initial expected dose value of *dose. CLOZAPINE*_(*t-*21),*i*_ for subject *i* at *t* =0; while the time-specific varying residual *e*_*ti*_ is the difference between the *dose. CLOZAPINE*_(*t-*21),*i*_ and the time-specific subject-mean value and it also represents the detrended value of *dose. CLOZAPINE*_(*t-*21),*i*_ for subject *i*. Therefore, the between-subject component *bz_dose. CLOZAPINE*_*i*_ is equivalent to *β*_0*i*_, and the within-subject component *wz_dose. CLOZAPINE*_(*t-*21),*i*_ is equivalent to *e*_*ti*_ .

### 2.6 Hybrid LMM analyses

#### 2.6.1 Model A

In Model A, a random intercept hybrid LMM model was built to identify significant associations between metabolic side effects and SGA medications. The model was defined as follows:

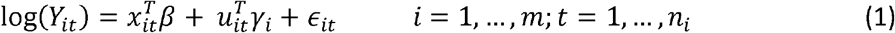

Here *Y*_*it*_ represents the metabolic measure of the *i*-th subject at *time t, n*_*i*_ denotes the number of measurements for the *i*-th subject, and *x*_*it*_ is the covariate vector for fixed effects (*β*), encompassing gender, education, age, treatment duration (*tx_duration*_*ti*_ ), between-subject medications (*bz.<drug>*), and within-subject medications (*wz.<drug>*). *u*_*it*_ is the covariate vector for random effects (*γ*_*i*_), set to 1 as Model A is a random intercept model. *ϵ*_*it*_ represents the error term. The subject ID serves as the grouping factor. The full formula of Model A expressed in *lmer4* (R package) format is stated in Supplementary Text 1.

#### 2.6.2 Model B

Hybrid LMM Model B, a random intercept model, maintained the structure of Equation (1), with the same covariate vectors *x*_*it*_ for fixed effects (*β*) and setting *u*_*it*_ to 1, similar to Model A. However, to estimate SGA-induced metabolic side effects per mg of antipsychotic drugs, the between- and with-subject medications were changed from binary variables (*bz.<drug>* and *wz.<drug>*) to continuous variables representing the dosages of the drugs (*bz_dose.<drug>* and *wz_dose.<drug>*). The full formula of Model B expressed in *lmer4* (R package) format is stated in Supplementary Text 2.

## 3. Results

Among our sample of 767 participants with SCZ, 46.7% (n = 358) were male, with a mean of 29.47 (SD = 9.17) years old. Participants’ mean education was 11.67 years (SD 2.90 years) and their mean onset age was 26.52 (SD 9.39). On average, participants received 5.5 (SD = 3.3) measures for lipid profiles (TC, HDL, LDL and TG), 5.5 (SD = 3.7) measures for BG, and 10.0 (SD = 9.0) measures for BMI. The mean number of prescription records for the entire sample was 180.7 per participant (SD = 72.9). Around 68% of our participants were drug-naïve at the time of baseline (pre-SGA) measures for the variables of interest. Among the SGA prescription records, olanzapine and aripiprazole were the most commonly prescribed SGAs in our cohort. The mean number of prescriptions for each SGA in our cohort is shown in Supplementary Table S2. To ensure the reliability of results, we scrutinized the homoscedasticity and normality assumptions for Model A and Model B (see Supplementary Text 3). The effect sizes presented below reflect the effects of specific SGAs, after controlling for other covariates, including other types of SGAs.

### 3.1 Metabolic side effects after SGA medications

Model A explored the associations between metabolic levels and SGA medications. Tables 1a and 1b highlight the estimated significance between-subject and within-subject associations (p < 0.05), whereas Table 2 lists the within-subject effect estimates.

**Table 1a.**
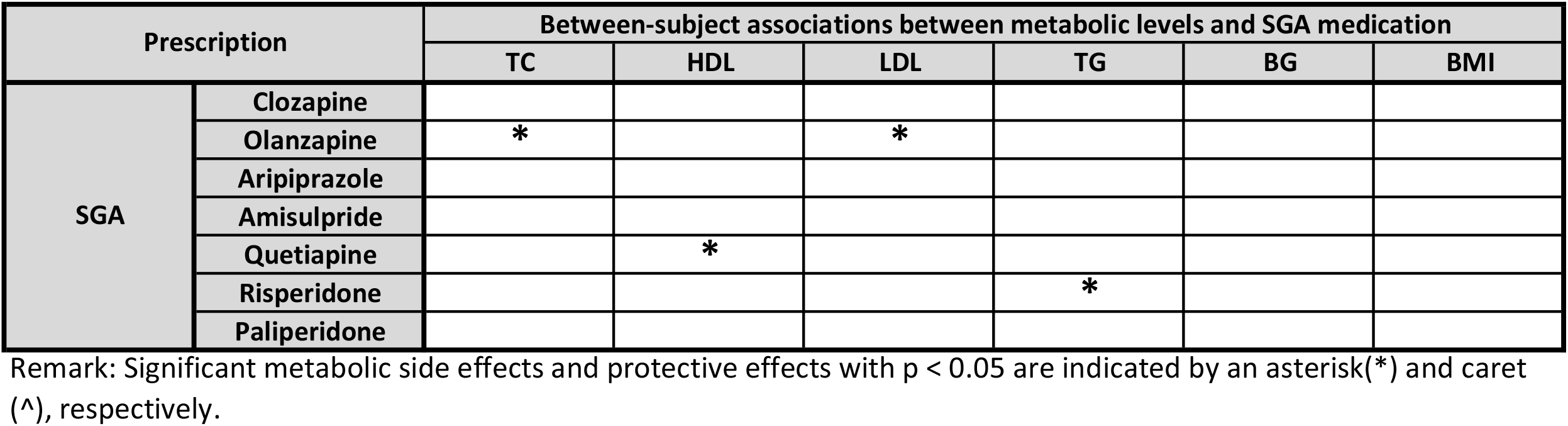
Between-subject associations between metabolic levels (or BMI) and SGAs with concomitant medications in Model A.

**Table 1b.**
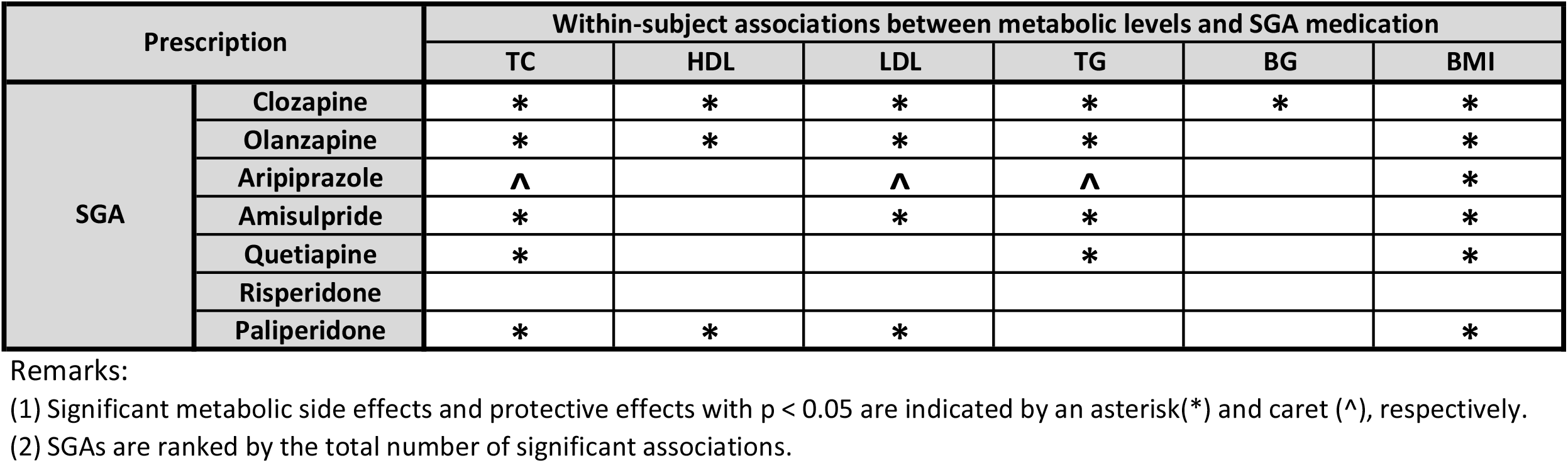
Within-subject associations between metabolic levels (or BMI) and SGAs with concomitant medications in Model A.

| Prescription |  | Within-subject associations between metabolic levels and SGA medication |  |  |  |  |  |
| --- | --- | --- | --- | --- | --- | --- | --- |
|  |  | TC | HDL | LDL | TG | BG | BMI |
| SGA | Clozapine | * | * | * | * | * | * |
|  | Olanzapine | * | * | * | * |  | * |
|  | Aripiprazole | ^ |  | ^ | ^ |  | * |
|  | Amisulpride | * |  | * | * |  | * |
|  | Quetiapine | * |  |  | * |  | * |
|  | Risperidone |  |  |  |  |  |  |
|  | Paliperidone | * | * | * |  |  | * |

- (1) Significant metabolic side effects and protective effects with $p < 0.05$ are indicated by an asterisk(\*) and caret (^), respectively. - (2) SGAs are ranked by the total number of significant associations.

**Table 1c.**
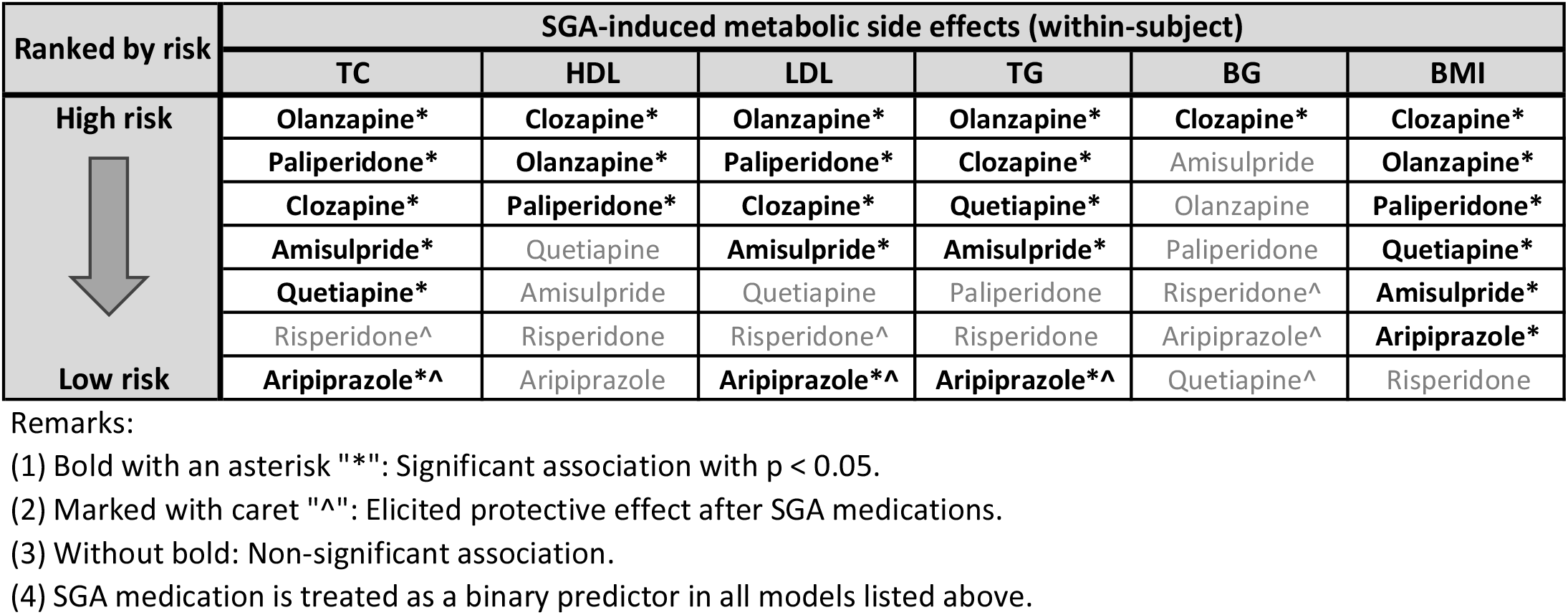
Severity of within-subject SGA-induced metabolic (or BMI) changes ranked by within-subject effect estimates listed in Table 2.

**Table 2.** Result of Model A - Within-subject effect estimates of SGAs with concomitant medications on the metabolic (or BMI) changes (n = 767).

| *Predictors* | TC |  |  | HDL |  |  | LDL |  |  | TG |  |  | BG |  |  | BMI |  |  |
| --- | --- | --- | --- | --- | --- | --- | --- | --- | --- | --- | --- | --- | --- | --- | --- | --- | --- | --- |
|  | *Estimates (SE)* | *t-value* | *p* | *Estimates (SE)* | *t-value* | *p* | *Estimates (SE)* | *t-value* | *p* | *Estimates (SE)* | *t-value* | *p* | *Estimates (SE)* | *t-value* | *p* | *Estimates (SE)* | *t-value* | *p* |
| wz.CLOZAPINE | 4.27 (0.06) | 3.049 | **0.002** | -7.9 (0.12) | 5.308 | **<0.001** | 8.53 (0.18) | 3.918 | **<0.001** | 21.39 (0.87) | 4.768 | **<0.001** | 8.6 (0.11) | 6.466 | **<0.001** | 5.9 (0.04) | 9.110 | **<0.001** |
| wz.OLANZAPINE | 6.92 (0.07) | 6.873 | **<0.001** | -5.24 (0.06) | 4.884 | **<0.001** | 9.09 (0.14) | 5.847 | **<0.001** | 25.11 (0.72) | 7.764 | **<0.001** | 0.85 (0.008) | 0.912 | 0.362 | 5.06 (0.02) | 12.673 | **<0.001** |
| wz.ARIPIRAZOLE | -3.81 (0.04) | 4.166 | **<0.001** | -0.01 (0.0001) | 0.010 | 0.992 | -4.11 (0.06) | 2.916 | **0.004** | -6.84 (0.19) | 2.560 | **0.01** | -1.31 (0.01) | 1.498 | 0.134 | 1.86 (0.007) | 4.890 | **<0.001** |
| wz.AMISULPRIDE | 3.72 (0.04) | 3.517 | **<0.001** | -1.69 (0.02) | 1.450 | 0.147 | 4.76 (0.08) | 2.917 | **0.004** | 9.99 (0.31) | 3.090 | **0.002** | 1.03 (0.01) | 1.037 | 0.3 | 2.65 (0.01) | 6.110 | **<0.001** |
| wz.QUETIAPINE | 3.01 (0.04) | 2.295 | **0.022** | -2.03 (0.03) | 1.405 | 0.16 | 2.82 (0.06) | 1.400 | 0.161 | 11.94 (0.46) | 2.943 | **0.003** | -2.36 (0.03) | 1.920 | 0.055 | 3.07 (0.02) | 5.245 | **<0.001** |
| wz.RISPERIDONE | -0.84 (0.01) | 0.597 | 0.551 | -0.1 (0.002) | 0.062 | 0.951 | -1.18 (0.03) | 0.547 | 0.585 | 2.11 (0.09) | 0.500 | 0.617 | -0.67 (0.009) | 0.510 | 0.61 | 0.4 (0.002) | 0.800 | 0.424 |
| wz.PALIPERIDONE | 4.47 (0.09) | 2.114 | **0.035** | -4.48 (0.1) | 1.960 | **0.05** | 8.87 (0.28) | 2.699 | **0.007** | 9.89 (0.61) | 1.538 | 0.124 | 0.81 (0.02) | 0.407 | 0.684 | 3.31 (0.03) | 3.875 | **<0.001** |

(1) The unit of effect estimates is the percentage changes in metabolic level (or BMI) after medication.
(2) SGA medication is treated as a binary predictor.

#### 3.1.1 Between-subject estimate

As expected, most associations between metabolic levels and between-subject SGA treatment effect did not reach statistical significance (see Table 1a), owing to confounding by indication/contraindication or other factors. However, several significant associations were found. Specifically, olanzapine was associated with an increase of 4.7% (95% CI: 4.5%,4.8%) in TC and an increase of 6.1% (95%CI: 5.8%, 6.5%) in LDL; quetiapine was associated with a decrease of 8.3% (95% CI: -8.9%, -7.7%) in HDL ; and risperidone was associated with an increase of 20.2% (95% CI: 17.3%, 23.1%) in TG.

Regarding the effects of demographics and duration of treatment (see Supplementary Table S3), our results showed that younger age was associated with better lipid profiles and BMI, and female SCZ participants showed better lipid profiles. A higher education level was associated with higher HDL and lower BG; but longer treatment duration was strongly associated with lower HDL and higher TG, BG, and BMI, but unexpectedly lower TC and LDL.

#### 3.1.2 Within-subject estimate

Most associations between metabolic levels and within-subject SGA treatment effects were significant (see Table 1b). Specifically, clozapine and olanzapine were strongly associated with worsened lipid profiles, BG and BMI, except the association between olanzapine and changes in BG failed to reach statistical significance. Aripiprazole was strongly associated with a reduction in TC of 3.8% (95% CI: -3.9%, -3.7%), a reduction in LDL of 4.1% (95%CI: - 4.2%, -4.0%), and a reduction in TG of 6.8% (95% CI: -7.2%, -6.5%), but an increase in BMI of 1.9% (95% CI: 1.8%, 1.9%). Amisulpride were associated with an increase in TC of 3.7% (95% CI: 3.6%, 3.8%), an increase in LDL of 4.8% (95% CI: 4.6%, 4.9%), an increase in TG of 10.0% (95% CI: 9.4%, 10.6%) and an increase in BMI of 2.7% (95% CI: 2.6%, 2.7%). Quetiapine was associated with an increase in TC of 3.0% (95% CI: 2.9%, 3.1%), an increase in TG of 11.9% (95% CI: 11.0%, 12.8%), and an increase in BMI of 3.1% (95% CI: 3.0%, 3.1%), but the association between quetiapine and changes in LDL failed to reach statistical significance. Paliperidone was associated with worsened TC (4.5%; 95% CI: 4.3%, 4.6%), HDL (-4.5%; 95% CI: -4.7%, -4.3%), LDL (8.9%; 95% CI: 8.3%, 9.4%) and BMI (3.3%; 95% CI: 3.3%, 3.4%). To further evaluate the impact of SGA medications on each metabolic parameter, the SGAs were ranked by their effect estimates in Table 1c.

### 3.2 Metabolic side effects at minimal effective dose of SGA medications

Model B estimated the dose-dependent within-subject estimate of SGAs on metabolic side effects. The within-subject percentage change in metabolic levels after taking one unit (mg) of the corresponding SGA per day is listed in Table 3, whereas the other effect estimates are listed in Supplementary Table S4. To enhance utility, we estimated the percentage changes of metabolic levels for each SGA, at the respective minimum effective dose (MED) used to manage SCZ patients (see Table 4a), based on linear dose-dependent relationship assumptions[41]. The MED was based on the Maudsley prescribing guidelines in psychiatry[42]. The SGAs were ranked from highest to lowest risk associated with metabolic side effects (without concerning the percentage changes). Among all the seven SGAs we studied, olanzapine showed the largest number of significant associations with metabolic side effects, while risperidone showed no significant associations. Aripiprazole showed a protective effect and was significantly associated with improving TC (-0.1740%/mg; 95%CI: - 0.1742%/mg, -0.1738%/mg) and TG (-0.4000%/mg; 95% CI: -0.4012%/mg, -0.3988%/mg). To further evaluate the impact of SGAs on each metabolic parameter, the SGAs were ranked by their percentage changes in metabolic levels after taking MED in Table 4b.

**Table 3.** Result of Model B - Dose-dependent within-subject effect estimates of SGAs with concomitant medications on the metabolic (or BMI) changes (n = 767).

| Predictors | TC |  |  | HDL |  |  | LDL |  |  | TG |  |  | BG |  |  | BMI |  |  |
| --- | --- | --- | --- | --- | --- | --- | --- | --- | --- | --- | --- | --- | --- | --- | --- | --- | --- | --- |
|  | Estimates (SE) | t-value | p | Estimates (SE) | t-value | p | Estimates (SE) | t-value | p | Estimates (SE) | t-value | p | Estimates (SE) | t-value | p | Estimates (SE) | t-value | p |
| wz_dose.CLOZAPINE | 0.008 (0) | 1.620 | 0.105 | -0.01 (0) | 2.363 | **0.024** | 0.02 (0) | 2.363 | **0.018** | 0.03 (0) | 2.078 | **0.038** | 0.01 (0) | 3.220 | **0.001** | 0 (0) | 0.122 | 0.903 |
| wz_dose.OLANZAPINE | 0.285 (0.0002) | 4.366 | **<0.001** | -0.18 (0.0001) | 4.196 | **0.013** | 0.42 (0.0004) | 4.196 | **<0.001** | 0.8 (0.002) | 4.161 | **<0.001** | 0.02 (0) | 0.398 | 0.691 | 0.18 (0) | 6.753 | **<0.001** |
| wz_dose.ARIPIRAZOLE | -0.174 (0.0001) | 3.464 | **0.001** | 0.03 (0) | 1.859 | 0.566 | -0.14 (0.0001) | 1.859 | 0.063 | -0.4 (0.0006) | 2.684 | **0.007** | -0.02 (0) | 0.518 | 0.604 | 0.04 (0) | 1.925 | 0.054 |
| wz_dose.AMISULPRIDE | 0.006 (0) | 2.662 | **0.008** | 0 (0) | 1.724 | 0.82 | 0.01 (0) | 1.724 | 0.085 | 0.02 (0) | 2.897 | **0.004** | 0 (0) | 0.029 | 0.977 | 0 (0) | 4.395 | **<0.001** |
| wz_dose.QUETIAPINE | 0.002 (0) | 0.483 | 0.629 | 0 (0) | 0.251 | 0.68 | 0 (0) | 0.251 | 0.802 | 0 (0) | 0.246 | 0.806 | -0.01 (0) | 1.445 | 0.148 | 0.01 (0) | 2.868 | **0.004** |
| wz_dose.RISPERIDONE | -0.495 (0.003) | 0.946 | 0.344 | 0.03 (0.0002) | 0.743 | 0.957 | -0.59 (0.0047) | 0.743 | 0.458 | -0.15 (0.0023) | 0.096 | 0.923 | -0.77 (0.004) | 1.570 | 0.116 | -0.07 (0.0001) | 0.382 | 0.702 |
| wz_dose.PALIPERIDONE | 0.636 (0.002) | 2.483 | **0.013** | -0.14 (0.0004) | 2.639 | 0.626 | 1.03 (0.004) | 2.639 | **0.008** | 0.9 (0.0068) | 1.196 | 0.232 | 0.37 (0.0009) | 1.579 | 0.114 | 0.35 (0.0004) | 3.034 | **0.002** |

Remark: The unit of effect estimates is the percentage change in metabolic level (or BMI) after taking one mg of drug per day.

**Table 4a.** Percentage changes of the metabolic level after taking a minimal effective dose (MED) of SGAs, estimated from the effect estimates listed in Table 3 and assuming a linear relationship between doses and SGA-induced metabolic changes.

| SGA | Minimal Effective Dose (MED) per day | Percentage change (within-subject) of metabolic level after taking one unit (i.e. one MED) of dose increment |  |  |  |  |  |
| --- | --- | --- | --- | --- | --- | --- | --- |
|  |  | TC | HDL | LDL | TG | BG | BMI |
| Olanzapine | 7.5 | 2.15 | -1.35 | 3.18 | 6.16 | 0.18 | 1.37 |
| Clozapine | 300 | 2.49 | -3.74 | 5.61 | 9.71 | 4.32 | -0.06 |
| Amisulpride | 400 | 2.55 | -0.24 | 2.51 | 8.41 | -0.04 | 1.82 |
| Paliperidone | 3 | 1.92 | -0.42 | 3.04 | 2.73 | 1.12 | 1.04 |
| Quetiapine | 300 | 0.69 | -0.66 | 0.54 | 1.03 | -1.93 | 1.54 |
| Risperidone | 4 | -1.97 | 0.13 | -2.34 | -0.59 | -3.03 | -0.27 |
| Aripiprazole | 10 | -1.73 | 0.32 | -1.43 | -3.92 | -0.24 | 0.41 |

- (1) The Minimal Effective Dose (MED) per day is adapted from The Maudsley Prescribing Guidelines in Psychiatry[39]. - (2) Bold: Significant association with $p < 0.05$ . - (3) SGAs are ranked by the total number of significant associations.

**Table 4b.** Severity of within-subject SGA-induced metabolic (or BMI) changes after taking a minimal effective dose (MED) of SGAs, ranked by percentage changes of metabolic level listed in Table 4a.

| Ranked by risk | SGA-induced metabolic side effects (within-subject) |  |  |  |  |  |
| --- | --- | --- | --- | --- | --- | --- |
|  | TC | HDL | LDL | TG | BG | BMI |
| High risk<br>↓<br>Low risk | Amisulpride* | Clozapine* | Clozapine* | Clozapine* | Clozapine* | Amisulpride* |
|  | Clozapine | Olanzapine* | Olanzapine* | Amisulpride* | Paliperidone | Quetiapine* |
|  | Olanzapine* | Quetiapine | Paliperidone* | Olanzapine* | Olanzapine | Olanzapine* |
|  | Paliperidone* | Paliperidone | Amisulpride | Paliperidone | Amisulpride^ | Paliperidone* |
|  | Quetiapine | Amisulpride | Quetiapine | Quetiapine | Aripiprazole^ | Aripiprazole* |
|  | Aripiprazole*^ | Risperidone^ | Aripiprazole^ | Risperidone^ | Quetiapine^ | Clozapine^ |
|  | Risperidone^ | Aripiprazole^ | Risperidone^ | Aripiprazole*^ | Risperidone^ | Risperidone^ |

- (1) Bold with an asterisk "\*": Significant association with $p < 0.05$ . - (2) Marked with caret "^": Elicited protective effect after SGA medications. - (3) Without bold: Non-significant association.

## 4. Discussion

Our study employed the hybrid LMM to account for the within- and the between-subject estimate of metabolic side effects of SGAs. However, many previous studies on SGA side effects did not distinguish between-subject averages from within-subject changes[9, 10] and thus might have reported inaccurate effect estimates, or inconsistent findings[7, 8, 43]. Our advanced statistical approach addressed this shortcoming of these previous studies and provided robust results regarding the long-term effects of SGAs on anthropometric and metabolic parameters in SCZ patients.

Although other observational studies have reported metabolic side effects of SGAs[44], our study was one of the most comprehensive and rigorous studies on the topic. Here we have studied a total of seven SGAs and covered six metabolic measures, enabling a comprehensive and systematic assessment of metabolic disturbances across almost all commonly used SGAs. Besides, as reported in the latest review[44], only few real-world studies have been conducted to investigate the effects of SGAs on weight gain or BMI changes, and very few reported quantitative changes in glucose or lipid levels (although there were a few studies on dyslipidaemia as a binary outcome). Very few real-world studies had included aripiprazole, and to our knowledge none included paliperidone. Furthermore, as we and other researchers have highlighted[44], confounding by indication is difficult to address in observational studies. To our knowledge, this is the first study to employ the within-subject approach to tackle this issue, which can account for unmeasured/unknown confounding factors that are time-invariant. Another strength is that we considered longitudinal changes of metabolic parameters using LMM, instead of only considering outcomes at the end of follow-up. The regular monitoring of metabolic profiles in our naturalistic cohort also reduced bias.

Our main findings are as follows. Regarding the within-subject estimate with SGAs as binary predictors (Model A), our findings suggested that most SGAs significantly altered metabolic profiles, aligning with a recent systematic review [8]. Specifically, risperidone did not show statistically significant metabolic side effects, consistent with another systematic review (except for BMI)[7]. Quetiapine had relatively modest metabolic side effects. Aripiprazole was associated with increased BMI but reduced TC, LDL, and TG. Clozapine, olanzapine and amisulpride worsened lipid profiles and BMI. Among the lipid measures, HDL was the least affected. In general, our findings concurred with earlier studies[7-10, 16, 43], after accounting for glucose- and lipid-lowering medications. Nevertheless, the results and effect sizes reported in this study may more accurately reflected the metabolic changes in SCZ patients over a longer duration (median ∼6.2 years), and the within-subject estimate may be closer to the true causal effect.

Of note, we observed that aripiprazole was associated with reduced TC, LDL and TG. Previous meta-analyses of RCTs mostly reported non-significant effects of aripiprazole on lipid profiles[7, 44]. However, several studies reported that when other SGAs were switched to aripiprazole or when aripiprazole was additional prescribed, patients were found to have improved lipid profiles [45-54]. These studies were in general limited by relatively modest sample sizes and short follow-up periods (longest = 52 weeks, others <=24 weeks), and none of the previous studies recruited a Chinese sample. Our findings may be considered as broadly consistent with these studies [45-54] which showed beneficial effects of aripiprazole in pragmatic settings. It should be noted that in our analysis of real-world SGA use, patients may be prescribed other SGAs with higher metabolic side effects before the use of aripiprazole, or have concomitant use of more than one SGA. The improved lipid profiles might happen in the context of an “already developed” hyperlipidemia after the use other SGAs.

In addition, we observed that paliperidone was associated with worsened metabolic profiles. Interestingly, we observed more significant metabolic side effects from paliperidone than from risperidone in this study. The novel finding requires further replication because the number of patients on paliperidone in our sample was relatively small.

Model B examined the dose-dependent effects of SGAs on metabolic parameters. At MED, olanzapine was associated with relatively wide-ranging side effects, i.e., worsening of 5 out of 6 metabolic parameters. It was followed by clozapine which was associated with 4 out of 6 metabolic parameters. On the other hand, clozapine induced the greatest percentage changes in HDL, LDL, TG, and BG; amisulpride had the largest effects on TC and BMI. Aripiprazole showed protective associations with TC and TG. These findings generally concurred with prior research[7, 8, 10, 16, 43], and may further inform clinicians about the SGA metabolic side effects at MED.

Across Model A (SGAs as binary predictor) and Model B (dosages as predictor; percentage change of metabolic parameters at MED being presented), we observed some subtle differences regarding the metabolic effects. When comparing the total number of significant associations between SGA and metabolic effects, clozapine ranked highest in Model A (see Table 1b), while olanzapine ranked first in Model B (see Table 4a). After dose adjustment at MED and comparing the percentage changes of metabolic levels after SGA medications, clozapine still ranked at the top (see Table 4b), but amisulpride and olanzapine had swapped positions (see Table 1c and Table 4b). Amisulpride may appear to have more severe metabolic side effects when compared with other SGA medications at MED, or when compared at higher dosages.

Our findings have important clinical implications. Given our more robust estimates of the SGA side effects in BMI and metabolic profiles in SCZ patients, clinicians and patients can be better informed for decision-making and side effect monitoring. They may also be better informed regarding the extent of metabolic abnormalities over a longer timeframe. To our knowledge, this is also the first real-world study on the effects of SGA dosage on metabolic parameters, which may provide a preliminary guide for clinicians. For example, the SGA doses for treating acute psychotic episodes and those for maintenance can differ. It may be useful to further study the optimal approach to adjust dosages considering the patient’s metabolic side effects to previous antipsychotic treatment. Our work provided novel data regarding the SGA side effects at MED and may help facilitate a more nuanced prescribing practice. Lastly, our finding that aripiprazole is associated with improved lipid profiles may also be of clinical interest.

### 4.1 Limitations

Our study has several limitations. First, we only focused on seven SGAs, yet some other SGAs were not covered. Second, lifestyle factors such as smoking and drinking, exercise, and diet were not measured and might have confounded our findings. Although the within-subject estimates are less susceptible to these confounders, residual confounding cannot be entirely ruled out in this observational study. Third, while our sample size provided over 19,316 observations due to the longitudinal design with multiple measurements, a larger sample size and longer follow-ups could strengthen the statistic power and precision of our findings. Additionally, the optimal response latency for metabolic changes induced by each SGA may differ, but we only chose the 21-day delay as the latency period for all SGAs, as informed by our modelling approach using AIC. Furthermore, our analysis of dosage effects assumed a linear relationship between dosage and metabolic parameters, which may be oversimplified. Further studies with larger samples and more advanced statistical methods are needed to explore potential non-linear dosage effects and confirm our findings.

### 4.2 Conclusion

Using a naturalistic longitudinal cohort design and sophisticated modelling to estimate within-subject effects, this study provides more robust estimates of the anthropometric and metabolic side effects of seven SGAs in Chinese SCZ patients. Clozapine and olanzapine were in general associated with the largest metabolic side effects. Aripiprazole was associated with protective effects against dyslipidemia. Our work may better inform both clinicians and patients of the heterogeneous metabolic risk profiles among SGAs. Consideration of these differing risk potentials can support optimal treatment decisions for individual patients.

## Data Availability

The data that support the findings of this study are available on request from the corresponding author. The data are not publicly available due to their containing information that could compromise the privacy of research participants.

## Contributions

**Kenneth C.Y. Wong**: Methodology, Software, Validation, Formal analysis, Writing – Original Draft, Visualization. **Perry B.M. Leung**: Resources, Data Curation. **Benedict K.W. Lee**: Resources, Data Curation. **Pak C. Sham**: Conceptualization, Methodology, Supervision, Writing - Review & Editing. **Simon S.Y. Lui**: Conceptualization, Methodology, Supervision, Writing - Review & Editing. **Hon-Cheong So**: Conceptualization, Methodology, Supervision, Writing - Review & Editing

## Acknowledgements

We thank all collaborators who were involved in patient recruitment and preparation of data, including Ms Hera Yeung, Ms Kirby Tsang, Dr Wong Ting Yat, Dr Karen Ho, Dr Karen Hung, Dr Eric Cheung and Dr KM Cheng.

## Supplementary Material

<u>Supplementary Tables</u>

<u>Supplementary Figures</u>

<u>Supplementary Text</u>

## Declaration of Generative AI and AI-assisted technologies in the writing process

During the preparation of this work the author(s) used Claude in order to edit this article for grammatical accuracy, and the original draft was written by the authors without the help of Claude. After using this tool/service, the author(s) reviewed and edited the content as needed and take(s) full responsibility for the content of the publication. Claude v2 was accessed from https://claude.ai/ and used without modification in November 2023.

## References

1. Correll, C.U., Current treatment options and emerging agents for schizophrenia. The Journal of Clinical Psychiatry, 2020. 81(3): p. 26548.

2. Grajales, D., V. Ferreira, and Á.M. Valverde, Second-generation antipsychotics and dysregulation of glucose metabolism: beyond weight gain. Cells, 2019. 8(11): p. 1336.

3. Turner, M.S. and D.W. Stewart, Review of the evidence for the long-term efficacy of atypical antipsychotic agents in the treatment of patients with schizophrenia and related psychoses. Journal of psychopharmacology, 2006. 20(6_suppl): p. 20–37.

4. Dong, M., et al., Prescription of antipsychotic and concomitant medications for adult Asian schizophrenia patients: findings of the 2016 Research on Asian Psychotropic Prescription Patterns (REAP) survey. Asian journal of psychiatry, 2019. 45: p. 74–80.

5. Davis, J.M., N. Chen, and I.D. Glick, A meta-analysis of the efficacy of second-generation antipsychotics. Archives of general psychiatry, 2003. 60(6): p. 553–564.

6. Lee, E., L.-Y. Chow, and C.-M. Leung, Metabolic profile of first and second generation antipsychotics among Chinese patients. Psychiatry research, 2011. 185(3): p. 456–458.

7. Pillinger, T., et al., Comparative effects of 18 antipsychotics on metabolic function in patients with schizophrenia, predictors of metabolic dysregulation, and association with psychopathology: a systematic review and network meta-analysis. The Lancet Psychiatry, 2020. 7(1): p. 64–77.

8. Pillinger, T., et al., Antidepressant and antipsychotic side-effects and personalised prescribing: a systematic review and digital tool development. The Lancet Psychiatry, 2023.

9. Zhou, T., et al., Weight changes following treatment with aripiprazole, risperidone and olanzapine: A 12-month study of first-episode schizophrenia patients in China. Asian Journal of Psychiatry, 2023. 84: p. 103594.

10. Zhang, Y., et al., Metabolic effects of 7 antipsychotics on patients with schizophrenia: a short-term, randomized, open-label, multicenter, pharmacologic trial. The Journal of Clinical Psychiatry, 2020. 81(3): p. 16879.

11. Mitchell, A.J., et al., Prevalence of metabolic syndrome and metabolic abnormalities in schizophrenia and related disorders—a systematic review and meta-analysis. Schizophrenia bulletin, 2013. 39(2): p. 306–318.

12. Rekhi, G., T.T. Khyne, and J. Lee, Estimating 10-year cardiovascular disease risk in Asian patients with schizophrenia. General Hospital Psychiatry, 2016. 43: p. 46–50.

13. Vancampfort, D., et al., Risk of metabolic syndrome and its components in people with schizophrenia and related psychotic disorders, bipolar disorder and major depressive disorder: a systematic review and meta-analysis. World psychiatry, 2015. 14(3): p. 339–347.

14. Laursen, T.M., T. Munk-Olsen, and M. Vestergaard, Life expectancy and cardiovascular mortality in persons with schizophrenia. Current opinion in psychiatry, 2012. 25(2): p. 83–88.

15. Castle, D.J. and E. Chung, Cardiometabolic comorbidities and life expectancy in people on medication for schizophrenia in Australia. Current Medical Research and Opinion, 2018. 34(4): p. 613–618.

16. Burschinski, A., et al., Metabolic side effects in persons with schizophrenia during mid- to long-term treatment with antipsychotics: a network meta-analysis of randomized controlled trials. World Psychiatry, 2023. 22(1): p. 116–128.

17. Mustafa, S., et al., Early stabilization of weight changes following treatment with olanzapine, risperidone, and aripiprazole: a 12-month naturalistic study of first episode psychosis. The Journal of Clinical Psychiatry, 2019. 80(5): p. 1087.

18. Vázquez-Bourgon, J., et al., Long-term metabolic effects of aripiprazole, ziprasidone and quetiapine: a pragmatic clinical trial in drug-naïve patients with a first-episode of non-affective psychosis. Psychopharmacology, 2018. 235(1): p. 245–255.

19. Li, S., et al., T4 and waist:hip ratio as biomarkers of antipsychotic-induced weight gain in Han Chinese inpatients with schizophrenia. Psychoneuroendocrinology, 2018. 88: p. 54–60.

20. Wu, X., et al., The comparison of glucose and lipid metabolism parameters in drug-naive, antipsychotic-treated, and antipsychotic discontinuation patients with schizophrenia. Neuropsychiatr Dis Treat, 2014. 10: p. 1361–8.

21. Bressington, D., et al., Cardiometabolic health, prescribed antipsychotics and health-related quality of life in people with schizophrenia-spectrum disorders: a cross-sectional study. BMC Psychiatry, 2016. 16(1): p. 411.

22. Curran, P.J. and D.J. Bauer, The disaggregation of within-person and between-person effects in longitudinal models of change. Annual review of psychology, 2011. 62: p. 583.

23. Eichler, H.-G., et al., Randomized Controlled Trials Versus Real World Evidence: Neither Magic Nor Myth. Clinical Pharmacology & Therapeutics, 2021. 109(5): p. 1212–1218.

24. Yang, S., et al., Development and validation of an age-sex-ethnicity-specific metabolic syndrome score in the Chinese adults. Nature Communications, 2023. 14(1): p. 6988.

25. Heymsfield, S.B., et al., Why are there race/ethnic differences in adult body mass index–adiposity relationships? A quantitative critical review. Obesity reviews, 2016. 17(3): p. 262–275.

26. Lear, S.A. and D. Gasevic, Ethnicity and metabolic syndrome: implications for assessment, management and prevention. Nutrients, 2019. 12(1): p. 15.

27. Zhang, R., et al., The Racial Disparities in the Epidemic of Metabolic Syndrome With Increased Age: A Study From 28,049 Chinese and American Adults. Frontiers in Public Health, 2022. 9.

28. Hamaker, E.L., Why researchers should think” within-person”: A paradigmatic rationale. 2012.

29. Joseph, K.S., A. Mehrabadi, and S. Lisonkova, Confounding by Indication and Related Concepts. Current Epidemiology Reports, 2014. 1(1): p. 1–8.

30. Twisk, J.W.R. and W. de Vente, Hybrid models were found to be very elegant to disentangle longitudinal within- and between-subject relationships. Journal of Clinical Epidemiology, 2019. 107: p. 66–70.

31. Allen, E., et al., Between- and within-subject associations of PTSD symptom clusters and marital functioning in military couples. Journal of Family Psychology, 2018. 32(1): p. 134–144.

32. Schirmbeck, F., et al., Stressful experiences affect the course of co-occurring obsessive-compulsive and psychotic symptoms: A focus on within-subject processes. Schizophrenia Research, 2020. 216: p. 69–76.

33. Schirmbeck, F., et al., Obsessive–compulsive symptoms in psychotic disorders: longitudinal associations of symptom clusters on between- and within-subject levels. European Archives of Psychiatry and Clinical Neuroscience, 2019. 269(2): p. 245–255.

34. Hirsch, L., et al., Second-Generation Antipsychotics and Metabolic Side Effects: A Systematic Review of Population-Based Studies. Drug Safety, 2017. 40(9): p. 771–781.

35. Lui, S.S., et al., A family study of endophenotypes for psychosis within an early intervention programme in Hong Kong: Rationale and preliminary findings. Chinese Science Bulletin, 2011. 56: p. 3394–3397.

36. VandenBerg, A.M., An update on recently approved long-acting injectable second-generation antipsychotics: Knowns and unknowns regarding their use. Mental Health Clinician, 2022. 12(5): p. 270–281.

37. Chávez-Castillo, M., et al., Metabolic risk in depression and treatment with selective serotonin reuptake inhibitors: are the metabolic syndrome and an increase in cardiovascular risk unavoidable. Vessel Plus, 2018. 2(6): p. 2574-1209.2018.

38. Kamarck, T.W., et al., Citalopram improves metabolic risk factors among high hostile adults: results of a placebo-controlled intervention. Psychoneuroendocrinology, 2011. 36(7): p. 1070–1079.

39. Rosenblat, J.D. and R.S. McIntyre, Pharmacological approaches to minimizing cardiometabolic side effects of mood stabilizing medications. Current Treatment Options in Psychiatry, 2017. 4: p. 319–332.

40. Fiorentino, N., et al., Metabolic Alterations and Drug Interactions: The Role of the Association between Antipsychotics/Mood Stabilizers and Cognitive Deficits. Psychiatria Danubina, 2022. 34(Suppl 8): p. 100–104.

41. Bishara, D., et al., Olanzapine: A Systematic Review and Meta-Regression of the Relationships Between Dose, Plasma Concentration, Receptor Occupancy, and Response. Journal of Clinical Psychopharmacology, 2013. 33(3).

42. Taylor, D.M., T.R. Barnes, and A.H. Young, The Maudsley prescribing guidelines in psychiatry. 2021: John Wiley & Sons.

43. Huhn, M., et al., Comparative efficacy and tolerability of 32 oral antipsychotics for the acute treatment of adults with multi-episode schizophrenia: a systematic review and network meta-analysis. The Lancet, 2019. 394(10202): p. 939–951.

44. Bernardo, M., et al., Real-World Data on the Adverse Metabolic Effects of Second-Generation Antipsychotics and Their Potential Determinants in Adult Patients: A Systematic Review of Population-Based Studies. Advances in Therapy, 2021. 38(5): p. 2491–2512.

45. Chen, Y., et al., Comparative effectiveness of switching antipsychotic drug treatment to aripiprazole or ziprasidone for improving metabolic profile and atherogenic dyslipidemia: a 12-month, prospective, open-label study. Journal of Psychopharmacology, 2012. 26(9): p. 1201–1210.

46. Fleischhacker, W.W., et al., Effects of adjunctive treatment with aripiprazole on body weight and clinical efficacy in schizophrenia patients treated with clozapine: a randomized, double-blind, placebo-controlled trial. International Journal of Neuropsychopharmacology, 2010. 13(8): p. 1115–1125.

47. Stroup, T.S., et al., A randomized trial examining the effectiveness of switching from olanzapine, quetiapine, or risperidone to aripiprazole to reduce metabolic risk: comparison of antipsychotics for metabolic problems (CAMP). American Journal of Psychiatry, 2011. 168(9): p. 947–956.

48. Wani, R.A., et al., Effects of switching from olanzapine to aripiprazole on the metabolic profiles of patients with schizophrenia and metabolic syndrome: a double-blind, randomized, open-label study. Neuropsychiatric disease and treatment, 2015: p. 685–693.

49. Fan, X., et al., Metabolic effects of adjunctive aripiprazole in clozapine-treated patients with schizophrenia. Acta psychiatrica scandinavica, 2013. 127(3): p. 217–226.

50. Newcomer, J.W., et al., A multicenter, randomized, double-blind study of the effects of aripiprazole in overweight subjects with schizophrenia or schizoaffective disorder switched from olanzapine. Journal of Clinical Psychiatry, 2008. 69(7): p. 1046–1056.

51. Takahashi, H., T. Oshimo, and J. Ishigooka, Efficacy and Tolerability of Aripiprazole in First-Episode Drug-Naive Patients With Schizophrenia: An Open-Label Trial. Clinical Neuropharmacology, 2009. 32(3): p. 149–150.

52. Henderson, D.C., et al., Aripiprazole added to overweight and obese olanzapine-treated schizophrenia patients. Journal of clinical psychopharmacology, 2009. 29(2): 53. p. 165–169.

53. Kim, S.-W., et al., Effectiveness of Switching to Aripiprazole From Atypical Antipsychotics in Patients With Schizophrenia. Clinical Neuropharmacology, 2009. 54. 32(5): p. 243–249.

54. De Hert, M., et al., A case series: evaluation of the metabolic safety of aripiprazole. Schizophrenia bulletin, 2007. 33(3): p. 823–830.

