## Supplementary Text for "Long-term Metabolic Side Effects of Second-Generation Antipsychotics in Chinese Patients with Schizophrenia: A Within-Subject Approach with modelling of dosage effects"

### Supplementary Text 1

The full formula of Model A expressed in lmer4 (R package) format is as follows.

Log(metabolic measure) ~ age + gender + education + tx_duration +

bz.CLOZAPINE + bz.OLANZAPINE + bz.ARIPIPRAZOLE + bz.AMISULPRIDE + bz.QUETIAPINE + bz.RISPERIDONE + bz.PALIPERIDONE +

bz.METFORMIN + bz.ATORVASTATIN + bz.SIMVASTATIN + bz.VALPROATE + bz.LITHIUM + bz.CITALOPRAM + bz.SERTRALINE +

wz.CLOZAPINE + wz.OLANZAPINE + wz.ARIPIPRAZOLE + wz.AMISULPRIDE + wz.QUETIAPINE + wz.RISPERIDONE + wz.PALIPERIDONE +

wz.METFORMIN + wz.ATORVASTATIN + wz.SIMVASTATIN + wz.VALPROATE + wz.LITHIUM + wz.CITALOPRAM + wz.SERTRALINE + (1|ID)

The *bz.<drug>* indicates the between-subject covariate of drug treatment, where 0 denotes no drug taken, and 1 indicates drug intake 21 days before the metabolic measurement. The *wz.<drug>* represents the within-subject covariate of drug treatment. The subject identifier (ID) serves as the grouping factor, denoted by (1|ID), indicating that Model A is a random intercept model.

### Supplementary Text 2

The full formula of Model B expressed in lmer4 (R package) format is as follows.

Log(metabolic measure) ~ age + gender + education + tx_duration +

bz_dose.CLOZAPINE + bz_ dose.OLANZAPINE + bz_dose.ARIPIPRAZOLE + bz_dose.AMISULPRIDE + bz_dose.QUETIAPINE + bz_dose.RISPERIDONE + bz_dose.PALIPERIDONE +

bz_dose.METFORMIN + bz_dose.ATORVASTATIN + bz_dose.SIMVASTATIN + bz_dose.VALPROATE + bz_dose.LITHIUM + bz_dose.CITALOPRAM + bz_dose.SERTRALINE +

wz_dose.CLOZAPINE + wz_dose.OLANZAPINE + wz_dose.ARIPIPRAZOLE + wz_dose.AMISULPRIDE + wz_dose.QUETIAPINE + wz_dose.RISPERIDONE + wz_dose.PALIPERIDONE +

wz_dose.METFORMIN + wz_dose.ATORVASTATIN + wz_dose.SIMVASTATIN + wz_dose.VALPROATE + wz_dose.LITHIUM + wz_dose.CITALOPRAM + wz_dose.SERTRALINE + (1|ID)

The *bz_dose.<drug>* represents the between-subject covariate of antipsychotic dose (mg), and *wz_dose.<drug>* is the within-subject covariate of antipsychotic dose (mg). Both covariates were recorded 21 days before the metabolic measurements. The subject identifier (ID) serves as the grouping factor, denoted by (1|ID), indicating that Model B is a random intercept model.

### Supplementary Text 3

#### Homoscedasticity and normality of residual in the Hybrid LMM models

Homoscedasticity and normality are standard assumptions in LMM. We scrutinized these assumptions for Model A and Model B to ensure the reliability of the results. As shown in Supplementary Fig. S3, residuals for both models exhibited a close-to-normal distribution, except for BG. Moreover, residual and fitted value plots for both models are shown in Supplementary Figs S4a and S4b respectively. The results suggested that homoscedasticity was generally upheld, except for a funnel shape in the BG plots, indicating a violation of the assumption of equal residual variance. Nevertheless, previous studies showed that linear mixed models are generally robust to violation of model assumptions[1].

### References:

1.	Schielzeth, H., et al., *Robustness of linear mixed-effects models to violations of distributional assumptions.* Methods in Ecology and Evolution, 2020. **11**(9): p. 1141-1152.
